# “I Been Taking Adderall Mixing it With Lean, Hope I Don’t Wake Up Out My Sleep”: Harnessing Twitter to Understand Nonmedical Prescription Stimulant Use among Black Women and Men Subscribers

**DOI:** 10.1101/2024.12.03.24318408

**Authors:** Joni-Leigh Webster, Sahithi Lakamana, Yao Ge, Abeed Sarker

## Abstract

Black women and men outpace other races for stimulant-involved overdose mortality despite lower lifetime use. Growth in mortality from prescription stimulant medications is increasing in tandem with prescribing patterns for these medications. We used Twitter to explore nonmedical prescription stimulant use (NMPSU) among Black women and men using emotion and sentiment analysis, and topic modeling. We applied the NRC Lexicon and VADER dictionary, and LDA topic modeling to examine feelings and themes in conversations about NMPSU by gender. We paid attention to the ability of natural language processing techniques to detect differences in emotion and sentiment among Black Twitter subscribers given increased mortality from stimulants. We found that, although emotion and sentiment outcomes match the directionality of emotions and sentiment observed (i.e., Black Twitter subscribers use more positive language in tweets), this belies limitations of NRC and VADER dictionaries to distinguish feelings for Black people. Even still, LDA topic models showcased the relevance of hip-hop, dependence on NMPSU, and recreational use as consequential to Black Twitter subscribers’ discussions. However, gender shaped the relevance of these topics for each group. Greater attention needs to be paid to how Black women and men use social media to discuss important topics like drug use. Natural language processing methods and social media research should include larger proportions of Black, Hispanic/Latinx, and American Indian populations in development of emotion and sentiment lexicons, otherwise outcomes regarding NMPSU will not be generalizable to populations writ large due to cultural differences in communication about drug use online.

## Introduction

Between 2007 and 2019, Black people experienced rapid growth in mortality from co-involvement of stimulants and opioids, with a 575 percent increase in death attributable to cocaine and opioids, and 16,200 percent increase in death involving methamphetamines and other stimulants and opioids (Townsend et al., 2022). Contamination of stimulant products with fentanyl and other synthetic opioids is driving rises in overdose deaths of Black women and men (Kline et al., 2023; Ciccarone, 2021; O’Donnell et al., 2020). Yet, mortality is not limited to Black adults, as Black people of all ages, including adolescents and young adults, demonstrate higher mortality from stimulants than White peers (Walia et al., 2021).

Illegalized stimulants like those previously mentioned are not the only culprits in mortality. Prescription medications like Adderall, used to treat attention disorders like ADHD, have increased stimulant-involved burdens of death among Black American and American Indians, outpacing White people who use drugs (CDC, 2023; Kariisa, 2021). Although diagnosis and treatment for ADHD have coincided with higher mortality rates, gaps in diagnosis and treatment of ADHD among Black people persists, with Black women and Black girls being less likely to be diagnosed with ADHD than White adults and children, and in some cases Black men (Bergey et al., 2022; Glasofer and Dingley, 2022; Shi et al., 2021; Fairman et al., 2021; Fairman et al., 2020; Chung et al., 2019). Underdiagnosis of ADHD can lead to self-medicating symptoms through nonmedical prescription stimulant use (the use of medications in ways not prescribed by medical providers or use of medications not prescribed to those taking them) for Black adolescents contributing to mortality among this group (Goodhines et al., 2020).

Like young people of all races, Black adolescents and young adults gain access to prescription stimulants through social networks including friends, family members and peers, and medical providers (Mendez et al., 2023; Fairman et al., 2021; Goodhines et al., 2020; Hulme et al., 2018). Of significance is the likelihood of exchanging one form of stimulant for another because of similar effects, like in the case of prescription stimulants and cocaine (Shearer et al., 2022). More research is needed to understand the role nonmedical prescription stimulant use (NMPSU) plays in mortality from stimulants given lower rates of use among Black adults (Shearer et al., 2022) and lack of evidence of demographic differences in patterns of use among Black adolescents in comparison to White peers (Goodhines et al., 2020). Twitter provides an opportunity for us to examine this in greater detail, as Black women and men use the site to communicate with one another and share experiences relevant to a Black cultural identity and to talk about mental health including drug use (Francis and Finn, 2021; Brock, 2020; Florini, 2014).

### The Implication of Race and Gender in Social Media Research on Nonmedical Drug Use

Black social media subscribers talk about NMPSU on Twitter more often than they discuss other forms of nonmedical prescription drug use (NMPDU; Yang et al., 2023). However, research examining the intersection of race and gender on drug use behaviors using social media data is limited. Meaning, while studies may account for gender and race in their sampling frames (Yang et al., 2023; Stevens et al., 2020), these variables are not used to observe differences in Twitter conversation topics about NMPDU. This is troubling since race and gender are related to drug behaviors because of experiences of racism, sexism, and other forms of oppression, as well as from socio-environmental risk factors like poverty and adverse childhood experiences which make NMPDU more likely (Banks et al, 2023; Silverstein et al., 2023; Gondre-Lewis et al., 2022; Nicholson, 2020; Miguel et al., 2019; Kramer et al., 2009; Link and Phelan, 1995).

In this way, race and gender intersect to create graduated effects on drug related outcomes among Black women and men (Crenshaw, 1991). Whereas women writ large are less likely than men to be given Narcan in the case of an overdose (Jones et al., 2023a), Black women are more likely to die from stimulant use than White women (Jones et al., 2023b). Even more, Black women and men experience differential dangers associated with exposure to violence at community and interpersonal levels, exacerbating the probability of testing positive for HIV and using stimulants to cope (Stockman et al., 2021; Nydegger et al., 2021; Saadatmand et al., 2020; Miguel et al., 2019; Lichtenstein, 2004; Crenshaw, 1991).

Nevertheless, race and gender are overlooked outside of survey methods and qualitative research (Nikunen, 2021). For example, despite impacting the possibility of diagnosis of and prescription of treatment for attention disorders due to provider bias, Guntunku et al. (2019) did not incorporate race or gender characteristics when examining how Twitter subscribers discussed ADHD. And, although Raza et al. (2023) provided a robust framework to investigate adverse events and symptoms of nonmedical use of Adderall and other prescription medications, they did not scrutinize the implications of race and gender on outcomes. Yet, evaluation of subscribers’ raced and gendered identities are not only important because of the impact of oppression on drug-related outcomes, but because these characteristics offer insight to how processes of racialization and gendering influence linguistic and thematic distinctions in conversations online (Nee et al., 2022).

In particular, race has been shown to affect communication patterns and conversations on Twitter (Preoţiuc-Pietro and Ungar, 2018) through cultural identity while gender shapes the kinds of emotional language used and topics discussed by women and men (Sun et al., 2020). Case in point, Black people on Twitter use African American English vernacular and other signifiers of a Black cultural identity frame to ensure safety when communicating with one another (Brock, 2020; Florini, 2014), which may impact Black people’s likeliness to disclose personal information, and to express positivity, anger, disgust, fear, and sadness on Twitter with one another in comparison to Hispanic/Latinx, Asian, and White subscribers (Preoţiuc-Pietro and Ungar, 2018). Blodgett et al. (2016) and Kiritchenko and Mohammad (2018) observed that lapses of inclusion of Black social media subscribers in digital research leads to biasing of NLP algorithms in the form of language classifiers and emotion/sentiment lexicons. This causes failures to detect Black Twitter subscribers’ speech patterns including syntactic abbreviations, grammar, and emotions that challenges the validity and interpretability of computational methods when they are included in social media research. This means that the collective gap in digital inquiry accounting for race and gender in social media research undermines the reliability of results by whitewashing outcomes and defaulting to a White man subscriber when developing NLP tools (Nee et al., 2022; Hampton, 2021; Noble, 2018).

By not attending to differences in communication patterns about drug-behaviors online and not observing the significance of gender and race in NLP techniques, social media research regarding drug-related harms is limited across populations. For instance, although Al-Garadi et al. (2021) used novel methods to disambiguate between types of prescription drug use chatter on Twitter as abuse, non-abuse consumption, mention only, and unrelated, they missed more granular differences regarding frequency of terms for each group (e.g., abuse versus non-abuse), as the terms “glass_whiskey” and “diesel_jeans” do not necessarily demonstrate personal abuse, but rather a commonly tweeted cultural artifact. Namely, in this context these words do not demonstrate co-consumption of drugs, but rather a reference to a popular YouTube video of a man consuming Adderall with whiskey while wearing diesel jeans. Delineating topics across groups could have showcased this discrepancy. In a similar way, while Stevens et al. (2020) accounted for race in their sampling frame and analytic methods, they did not report the implication of race on results regarding drug-related keywords and topics. Though they used “slang” terms and song lyrics to construct classifiers, they did not inspect their effects at inter-group levels to understand Black and White distinctions in drug-behaviors and communication patterns on Twitter.

Taken together, Black women and men are likely to communicate similarly to one another in terms of cultural ascriptions tied to race, and distinctly when gender affects emotional expressions and topics discussed. We bridged the gaps of previous studies by investigating the intersection of race and gender in greater detail as it concerns NMPSU, because Black women and men have different experiences tied to NMPSU than other racial groups. We focused on emotion and topic analysis specifically, because Al-Garadi et al. (2022) showed gendered differences wherein women Twitter subscribers were more likely than men to express positive emotions, anticipation, sadness, and joy in comparison to men, who were more likely to express feelings of anger in tweets about NMPSU.

## Methods

### Data Collection

We augmented work by Raza et al. (2023), and Al-Garadi et al. (2021, 2022) to determine how Black Twitter subscribers talk about nonmedical prescription stimulant use, and how gender shaped these conversations. Tweets were collected from March 6, 2018, to April 30, 2021 using keyword-based pipelines to extract gender and race from Twitter profile content, with race assigned by self-identification in historical tweets as someone who is Black (e.g., “I am Black”; Yang et al. 2023). Pipelines produced high levels of predictive accuracy for gender and race (Yang et al., 2023; Yang et al., 2021). We extracted approximately 2,418 tweets from women and 2,702 tweets from men from a total of 1,506 Black women and 1,703 Black men Twitter subscribers (Table 1). All tweets were publicly available, and usernames were removed to maintain privacy.

**Table 1.**
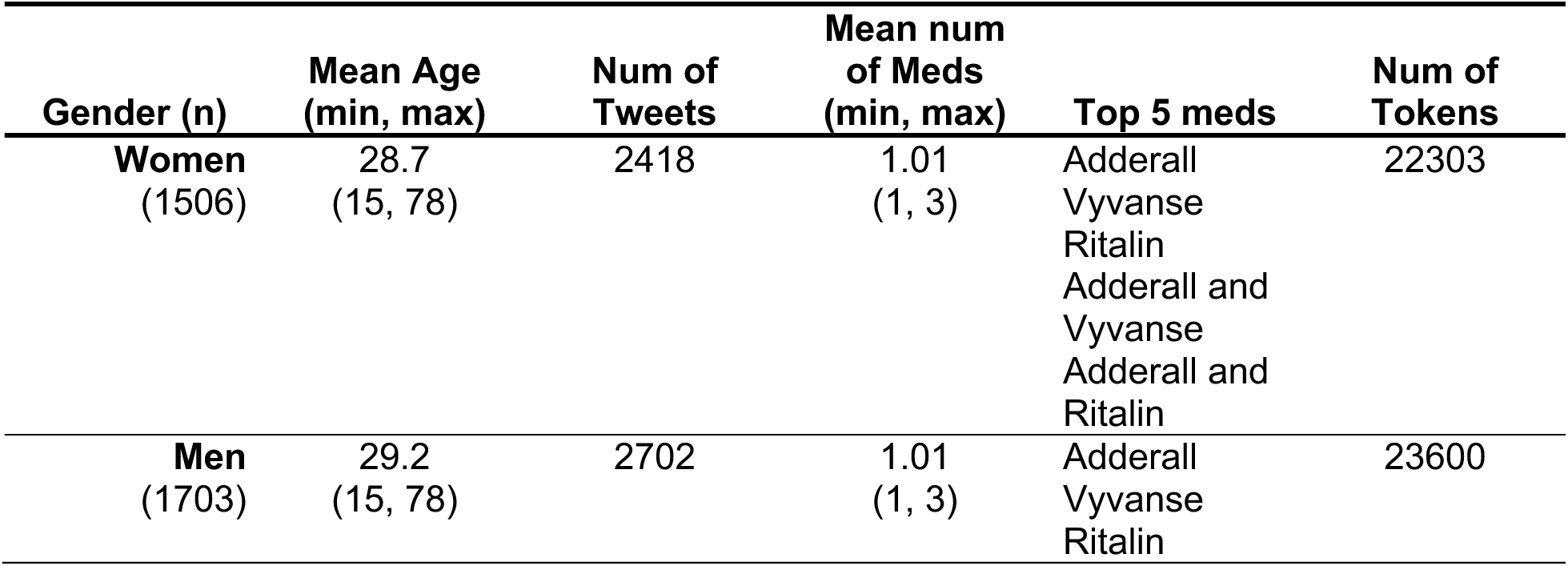
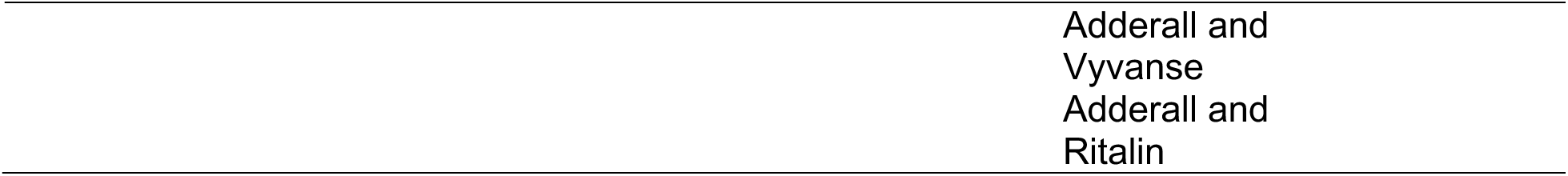
Black Twitter Subscriber Tweet Distribution.

### Analytic Methods

#### Emotion Analysis

We conducted emotion analysis using the NRC lexicon, which uses English words to identify the emotional content of text depicting feelings of anger, anticipation, disgust, fear, joy sadness, surprise, and trust, as well as their positive and negative valence (Mohammad and Turney, 2013; Plutchik, 2001). Values for each emotion were dependent on their proportion representation across each tweet. However, we removed tweets assigned a value of 0 for all emotions, indicating an inability of NRC to detect emotions overall, reducing the number of tweets in our sample to 1,631 tweets for women and 1,755 tweets for men. We did this because emotion and sentiment dictionaries fail to distinguish African American English vernacular and syntax for Black versus White Twitter subscribers, and to increase accuracy of our comparative assessments by gender (Blodgett et al., 2016). We used median values for each emotion to exclude outliers, leaving approximately 440 tweets for Black women Twitter subscribers and 436 tweets for Black men subscribers. The Mann-Whitney U Test helped us identify statistically significant differences in emotions expressed by women and men considering non-normality of data (Table 2; McKnight and Najab, 2010; Nachar, 2008).

**Table 2.** NRC Lexicon Emotion Analysis.

| Emotions | Median: Women | Median: Men | Mann-Whitney U Test |  |
| --- | --- | --- | --- | --- |
|  |  |  | p-value (p) | Effect size (r) |
| **Positive** | 0.222 | 0.2 | 0.206 | -0.0427 |
| **Negative** | 0.167 | 0.2 | 0.0634 | 0.0627 |
| **Anger** | 0 | 0 | 0.851 | 0.00636 |
| **Anticipation** | 0.0646 | 0.0833 | 0.702 | 0.0129 |
| **Disgust** | 0 | 0 | 0.452 | -0.0254 |
| **Fear** | 0 | 0.0833 | 0.422 | 0.0271 |
| **Joy** | 0 | 0 | 0.531 | -0.0212 |
| **Sadness** | 0 | 0 | 0.568 | -0.0193 |
| **Surprise** | 0 | 0 | 1 | 0 |
| **Trust** | 0.125 | 0.111 | 0.456 | -0.0252 |

Size effects(r): \*Small difference ( $0.1 \leq r < 0.3$ ). \*\*Medium difference ( $0.3 \leq r < 0.5$ ). \*\*\*Large difference ( $0.5 \leq r < 0.7$ ). \*\*\*\*Very large difference ( $r \geq 0.7$ )

#### Sentiment Analysis

To account for potential accuracy limits of the NRC Lexicon emotion analysis, we supplemented these methods using the VADER sentiment dictionary, which determines the degree to which language in text is considered positive, negative, or neutral, assigning a compound score to detect polarity (Czarnek and Stillwell, 2022; Ribeiro et al., 2016; Hutto and Gilbert 2014; Table 3). Whereas positive, negative, and neutral scores can range between 0 and 1, compound scores are between -1 and 1, with neutrality assigned to values -0.05 to 0.05. We used the original sample o tweets by Black women and men subscribers who talked about NMPSU and removed those with compound scores showing high degree of neutrality (score of 0), because this signified difficulty assigning sentiment to each tweet. As such our tweet sample was reduced for women and men, with 1,597 and 1,665 tweets remaining, respectively. We followed the same procedures from our emotion analysis and conducted Mann-Whitney U Tests for each sentiment to determine gender differences in sentiment valence for Black women and men Twitter subscribers following the removal of outliers (final tweet sample of 1,544 tweets for women and 1,630 tweets for men).

**Table 3.** VADER Sentiment Analysis.

| Sentiment | Median: Women | Median: Men | Mann-Whitney U Test |  |
| --- | --- | --- | --- | --- |
|  |  |  | p-value (p) | Effect size (r) |
| **Positive** | 0.12 | 0.122 | 0.794 | 0.00463 |
| **Negative** | 0.095 | 0.087 | 0.493 | 0.0122 |
| **Neutral** | 0.77 | 0.776 | 0.239 | -0.0209 |
| **Compound** | 0.202 | 0.153 | 0.346 | -0.0167 |

*Size effects(r): \*Small difference ( $0.1 \leq r < 0.3$ ). \*\*Medium difference ( $0.3 \leq r < 0.5$ ). \*\*\*Large difference ( $0.5 \leq r < 0.7$ ). \*\*\*\*Very large difference ( $r \geq 0.7$ )*

#### Topic Models

For topic modeling, we used LDA (Latent Dirichlet Allocation), a probability-based model that assigns words to topics based on the likelihood of co-occurrence across documents (Blei et al., 2003). This is an unsupervised approach, which does not incorporate annotation methods, dictionaries, or data categorization to predict relevant topics. We assessed topic models for a range of 5 to 40 topics for each group (Black women and Black men), and inspected coherence values for each model. Coherence measures are used to identify how much words in a topic are related. We chose models with higher coherence values by gender and located the five-most prevalent topics across each model. Finally, we selected a 40-topic model for women and 30-topic model for men using the average percent distribution (minimum of 60 percent mean contribution) of prominent topics per tweet. We did this to ensure appropriate labeling of data, as coherence scores along with topic relevance, and word exclusivity, are important to model interpretation while considering gendered differences in topic representation (Tables 4-5; Weston et al., 2023; Yoon et al., 2013). We present labels before and after manual inspection of tweets alongside sample tweets to demonstrate the utility of qualitative inquiry in natural language processing to evaluate topic models of understudied populations. We observed 10 tweets per topic (when possible) with the highest percentage contribution to ensure themes closely reflected topics. Some topics were given multiple labels because the same word combinations could be used to describe various aspects of NMPSU.

**Table 4.** Women’s Most Prominent Topics (40-Topic Model)

| Topic Number | Topic Ngrams | Label (Before Annotation) | Label (After Annotation) | Sample Tweet |
| --- | --- | --- | --- | --- |
| 17 | take; need; adderall; drink; full; bottle; feel_like; rn; feel; thank | Cravings | Thoughts/feelings about stimulants (including cravings) | "I need more adderall or another brain" |
| 0 | day; adderall; think; give; god; every; fun; white; addiction; accidentally | Addiction/dependence | Addiction/Dependence | "@subscriber yea...it was an addy day #adderalldon [crying laughing emoji]" |
| 1 | like; know; adderall; never; drug; said; let; asked; girl; need | Experimentation | Effects of prescription stimulant use (includes noticing effects on others or others noticing the effects on the individual who consumed) | "what I look like after binging some adderall" |
| 4 | feel_like; feel; two; like; adderall; last_night; tomorrow; used; night; last | How use affects the day (e.g., at night, next day, etc.) | Storytelling about NMPSU (including based on time of day) | "getting drunk and on addies feels like youre stupid but faster" |
| 32 | much; coffee; adderall; taking; extra; lot; getting; please; ritalin; ive | Taking higher dose or supplementing prescription stimulants w/ caffeine | Negative effects of NMPSU (side effects from consuming large quantities or with caffeine) | "vitamin r - crushed. kmfdm - on repeat. coffee - extra strong. hydros - why not." |

**Table 5.** Men’s Most Prominent Topics (30-Topic Model)

| Topic Number | Topic Ngrams | Label (Before Annotation) | Label (After Annotation) | Sample Tweet |
| --- | --- | --- | --- | --- |
| 8 | taking_adderall; adderall; taking; feel; mixing; better; mixing_lean; eat; snorted; tonight | YB NBA - Genie lyrics | YB NBA - Genie lyrics | "I'm taking adderall mixing it with lean hope I don't wake out my sleep" |
| 19 | get; day; work; adderall; know; want; caffeine; damn; three; 14 | Stimulant use to get through the day (including caffeine) | Access or source | "someone bring me an adderall to my job. I will pay you \$\$\$" |
| 11 | take; back;<br>adderrall; make;<br>study; lot; made;<br>I'll; tweet; good;<br>like | Distracted from<br>studying (side<br>effect) | Dependence on<br>stimulants (and<br>cognitive side<br>effects) | "a solid 80% of<br>my tweets are<br>while under the<br>influence of addy.<br>My bad" |
| 22 | adderrall; really;<br>sleep; f***; give;<br>school; good;<br>play; call;<br>year_old | Stimulant use for<br>different activities<br>including<br>schoolwork | Lil Wayne ft.<br>Takeoff - I Don't<br>Sleep | "I don't sleep...f***<br>a bed...f*** a<br>spread...f*** a<br>sheet...f*** a<br>xan...give me<br>adderrall and<br>weed" |
| 14 | like; adderrall;<br>weed; year; help;<br>cup; pop; house;<br>beer; yet | Polydrug use | Recreational<br>use/information<br>about drug<br>experiences | "please god can I<br>find adderrall on<br>the floor again like<br>the day before" |

All assessments were conducted using Python programming language and standard preprocessing techniques (i.e., lowercasing; removal of stop words, hyperlinks, and phone numbers; lemmatization). We present modified versions of tweets to ensure subscriber confidentiality due to the sensitive nature of our study.

## Results

The mean age of Black women and men Twitter subscribers who engaged in NMPSU rounded to 29 years for both groups (Table 1), showing comparable representation between our larger study and Twitter subscribers who talk about nonmedical prescription stimulant use on the site (Raza et al., 2023; Yang et al., 2023). Women and men used the same mean number of stimulant medications (1.01 with a standard deviation between 1,3). Adderall followed by Vyvanse, and Ritalin were the most common medications in our dataset across groups. However, compared to men, women were slightly more likely to tweet about nonmedical prescription stimulant use based on proportion of tweets by gender. This reflects differences in posting patterns by which Black women post Twitter messages more often than Black men, instead of representing differential rates of use by gender (Bailey, 2021; Hargittai 2020).

### Emotion Analysis

In comparison to men, median proportion of tweets dedicated to expressing positive emotions and trust were higher for women (0.222 and 0.125 for women and 0.200 and 0.111 for men; Table 2). Whereas those describing negative, anticipatory, or fear-based emotions were greater for men (0.200, 0.0833, 0.0833 for men versus 0.167, 0.0646, 0.000 for women, respectively). Despite this, Mann-Whitney U Tests did not detect differences in emotion scores for women and men at significance levels of 0.05 or less. Though, there was a marginally significant difference in negative emotion expression for men versus women, with men being more likely to describe negative feelings in relation to NMPSU in their tweets (p = 0.0634). The effect size of this was negligible.

### Sentiment Analysis

Using the VADER sentiment analyzer, women and men’s tweets reflected similar proportions of positive, negative, and neutral language, with greatest proportion of tweets highlighting neutral, followed by positive, and negative sentiment (neutral = 0.770 vs 0.776; positive = 0.120 vs 0.122; negative = 0.0950 vs 0.0870 for women and men, respectively; Table 3). Although, compound scores were higher for women (0.202 versus 0.153), indicating slightly more positive language in women’s tweets, matching NRC results about use of more positive language by women. Like Mann-Whitney results for the NRC Lexicon, differences in valence of scores for Black women and men for all sentiments were not significant. Effect size scores mirrored these results.

### Topic Models

Moving to topic models, we found gendered differences in themes discussed by Black women and men in relation to nonmedical prescription stimulant use. Exploring women’s tweets first, we found that the most prevalent topics in order of most to least predominant were labeled for cravings; addiction/dependence; experimentation; how NMPSU affects the subscriber’s day or how time determines NMPSU; and taking higher dose of prescription medications or supplementing prescription stimulants with caffeine (Table 4). Labels prior to annotation corresponded to those after reviewing tweets, except for the label for experimentation. Experimentation contained “like”, “know”, “Adderall”, “never”, “drug, “said”, “let”, “asked”, “girl”, and “need” as dominant words, which we inferred to indicate willingness to try prescription stimulants because of “like”, “never”, “drug”, and “asked”. However, further inspection showed that these tweets were about the effects of prescription stimulant use, including noticing the effects NMPSU on others or others noticing the effects of NMPSU on the Twitter subscriber.

Unlike posts by women, we had more difficulty correctly labeling tweets for men. We mislabeled prominent themes for 3 out of 5 topics. Whereas women’s tweets were about experiences of NMPSU or dependence, men talked about pop culture via song lyrics along with accessing prescription stimulants or finding a source, and recreational use or using Twitter for information seeking about NMPSU (Table 5). For instance, we coded topic 22 for men as NMPSU for different activities including schoolwork. But, upon reading tweets linked to the topic we realized that Black men were in fact quoting lyrics from a song by Lil’ Wayne featuring Takeoff, “I Don’t Sleep”. The topic words were “Adderall”, “really, “sleep”, “f***”, “give”, “school”, “good”, “play”, “call”, and “year old”. The song lyrics are “F*** a bed, f*** a spread, f*** a sheet (I don’t sleep), f*** a Xanny, give me Adderall and weed (I don’t sleep)”. Our original label was not far off from the content of lyrics which describe NMPSU for various activities like sex, productivity towards reaching goals, and recreational use. However, we were unable to infer whether song lyric content matched Black men’s NMPSU patterns in relation to the topic identified. Conversely, we were able to recognize lyrics of YoungBoy Never Broke Again for topic 8 matching the song, “Genie”. Based on these lyrics, Black men expressed passive suicidal ideation in relation to consumption of prescription stimulants without providing context as to what might trigger listing these kinds of lyrics in tweets.

## Discussion

This study examined the utility of natural language processing (NLP) techniques like emotion analysis, sentiment analysis, and topic modeling to identify race- and gender-based patterns of NMPSU among Black women and men Twitter subscribers. Our results indicate that while NLP methods offer a way to examine large swathes of social media content, outputs are not generalizable across populations and require further inspection to ensure accurate interpretation of results (El-Azab and Nong, 2023). For instance, although the lead author (JW) identifies as a Black woman, who is a Twitter subscriber, her gender identity as a woman limited her ability to detect gendered differences in conversations about NMPSU for Black men, including topics about hip-hop as a relevant cultural signifier in Black men’s tweets. While other research notes the prevalence of song lyrics in tweets, they are usually discarded and not considered to reflect personal instances of drug use in social media research (Kim et al., 2020; Preotiuc-Pietro and Ungar, 2018). We recommend against this, since music can provide domain level knowledge not easily recognizable to researchers wherein subgroups of people who engage in nonmedical prescription drug use purposely invoke lyrics to denote subjective experiences tied to use (Semenza, 2018). For example, the invocation of hip-hop songs when talking about suicide and thoughts of overdose were reflected in tweets by Black men who use opioids (Webster, 2024). This indicates that music, especially hip-hop, is not just a marker of identity (Roy and Dowd, 2010), but a tool by which Black people seek safety and peace when struggling with mental health (Wright, 2019; Harper and Jackson, 2018; DeNora, 2013). Even more, there may be intergroup differences where those racialized as LatinX, White, or American Indian quote lyrics or mention media intrinsic to their own cultural repertoire, which is not consonant with groups racialized in other ways. Like in the case of hip-hop music and Black men Twitter subscribers.

Whereas past studies show gender-based differences in emotional expression for women and men who engage in NMPSU (Al-Garadi et al., 2022), we did not find this despite pulling data from the same corpus. This may be due to us exclusively analyzing tweets by Black Twitter subscribers whose gender could be predicted. As a result, using these characteristics to categorize data showed the limitations of current emotion and sentiment lexicons, like the NRC Lexicon and VADER dictionary, to detect emotion and sentiment for subscriber tweets since studies accounting for gender still default to White men social media subscribers. It is possible that greater representation of tweets by White women and men in training models used to create these dictionaries obfuscates results at subgroup levels, leading to an inability to elucidate cultural distinctions in linguistic expressions for Black women and men Twitter subscribers (Kiritchenko and Mohammad, 2018).

Though language is not a barrier in dictionary-based emotion analysis of our tweets, as all tweets were in English, NRC and VADER failed to detect the emotions and sentiments of more than 30 percent of tweets, with NRC demonstrating more restrictions. Even still, there were inconsistencies in which tweets were likely to be rated as neutral or showing no detectable emotion across dictionaries. Meaning, VADER tweets showing no sentiment were not always similarly assessed by NRC or vice versa. However, this could be due to differences in the words used by each lexicon, as NRC has a more robust dictionary (Al-Garadi et al., 2022).

At closer study by sampling and reviewing missing tweets by dictionaries, we found that tweets about time did not show sentiment distinctions for VADER, and that colloquial patterns of speech impeded the NRC lexicon from detecting emotions (per the assessment of JW). Further, among tweets removed for emotion and sentiment scores across [both] dictionaries, we noticed that they concentrated on language about need and functioning in relation to NMPSU, substantiating our claims regarding the limitation of dictionaries to assess group-level patterns in speech across racialized and gendered populations.

Still, topic models revealed gendered differences in nonmedical prescription stimulant use behaviors for Black women and men Twitter subscribers. While both groups talked about negative side effects from use, like cognitive impairment. The most prominent topics among women were related to dependency. Women were more likely to talk about craving prescription stimulants or relying on NMPSU to meet their needs, with Black women describing taking higher amounts of prescription stimulants and co-using stimulants with caffeine. This aligns with reports of women moving more quickly towards dependence than men (Bobzean et al., 2014). However, this does not mean that prescription stimulant dependance was not relevant to Black men since it was also among their most prevalent topics. Yet, unlike Black women, Black men were more likely to quote song lyrics about NMPSU, and to talk about issues accessing or finding sources for prescription stimulants, as well as engaging in information seeking about NMPSU on Twitter.

Hip-hop was significant in Black men’s conversations about NMPSU, as song lyrics from artists like YoungBoy Never Broke Again and Lil Wayne were featured in two out of five of men’s most prominent tweet topics. Future studies should evaluate how pop culture shapes or informs nonmedical prescription stimulant use among Black women and men since they display different degrees of influence in tweet content. Although Black women were less likely to include music in tweets, trigrams (word combinations that exist in context of text documents) ranked by gender show that cultural markers about hip-hop, comedy, and popular online videos were important to Black women’s conversations about NMPSU (Supplementary Tables & Figures: 1-5).

Since hip-hop as a genre is associated with galvanizing online discussions related to mental health for Black social media subscribers, computational studies should incorporate hip-hop to analyze the co-occurrence of mental health disorders among people who use drugs (Banks et al., 2023; Cenat et al., 2023; Raza et al., 2023; Francis 2021, Francis 2018). For instance, rises in conversations about suicide and mental health on Twitter and higher call volume to the National Suicide Prevention Lifeline followed the release of Logic’s song, “1-800-273-8255”, about seeking help when struggling with suicidal ideation (Niederkrotenthaler et al., 2021; Torgerson et al., 2021). While these studies did not account for race-based differences in Twitter conversations linking mental health to hip-hop, Francis (2021, 2018) showed that Black men discussed mental health and depression on Twitter following rapper Kid Cudi’s disclosure of suicidal ideation and depression on Facebook, with Black men being more likely to seek help about their depressive symptoms. This supports topic 8 of our model about Black men’s tweets, wherein, YoungBoy Never Broke Again’s lyrics from “Genie” describe passive suicidal ideation, and a willingness to mix prescription stimulants with lean (a candy and soda mixture with opioids, like liquid codeine, and promethazine) to die. We do not believe this is innocuous since listing lyrics in tweets coincides with the presentation of mental health concerns for conditions like depression, and content about mental health struggles are rife in hip-hop songs (Alavijeh et al., 2023; Kresovich et al., 2021; Wright, 2019; Harper and Jackson, 2018; Bailey, 2013). Moreover, Al-Garadi infer the words “taking mixing” are about dosage in NMPDU tweets, however, we hypothesize they reflect the song by YoungBoy Never Broke Again we described above.

### Limitations

Although our study advances research on drug related outcomes using Twitter to account for how race and gender, as fundamental causes of disease, shape health, there are limitations. For instance, our corpus contained a little more than 5,000 tweets by Black Twitter subscribers who engaged in NMPSU. A larger sample may account for discrepancies in detecting emotion and sentiment across tweets, as posts in our sample may not be generalizable to all Black Twitter subscribers who engage in NMPSU. Even still, we do not believe this is the case since investigations of sentiment dictionaries display racialized disparities in detecting emotions, with stereotypical emotions assigned to Black social media subscribers (Kiritchenko and Mohammad, 2018) and an undercounting of depressive symptom words for Black Facebook subscribers in comparison to White (Rai et al., 2024). Moreover, topics detected in our analyses match those of Raza et al. (2023) which investigated NMPSU and identified topics about mental health, side effects, and addiction/dependence.

Although a strength of our study is that we focus on the implications of race and gender on stimulant use, we do not assess differences across racial groups or across drug categories (i.e., opioids, benzodiazepines, polydrug use) for nonmedical prescription drug use. Because race is inferred, we were unable to determine whether all subscribers in our repository self-identified with a specific race, impeding our ability to predict races for all subscribers in the full dataset at subgroup levels. As a result, race-based differences for drug behavior patterns among Hispanic/Latinx, Asian, and American Indian populations were limited because of lower volume of tweet representation in our dataset. More work is needed to detect tweets for Black, Hispanic/Latinx, and American Indian populations given rising rates of opioid- and stimulant-involved overdose deaths among these groups, and because their experiences are rarely investigated (Kariisa et al., 2022; Kariisa et al., 2021).

## Conclusion

To our knowledge our study is the first to use Twitter to investigate how race and gender intersect in meaningful ways to shape drug-related discussions online using natural language processing techniques, with special attention paid to Black Twitter subscribers. We find that Twitter offers a space for public health researchers in the digital domain to delve more deeply into conversations about nonmedical prescription stimulant use among Black people who use drugs, enabling them to evaluate whether topics relevant to subscribers on the site match real world drug-related experiences more broadly. We believe understanding group level differences can create opportunities for targeted interventions to mitigate drug-related harms through social media, like in the case of Twitter discussions spurred by rappers Kid Cudi and Logic in relation to suicide and depression.

Though our results show less robust emotion and sentiment detection than Al-Garadi et al. (2022), we believe the inclusion of Black Twitter subscribers as part of analytic teams can improve NLP methods to augment current lexicons by providing domain specific knowledge of African American vernacular and culture, and its significance in shaping syntax and conversations online (i.e., Black Twitter as a digital community and cultural phenomenon; Aguilar et al., 2023; Frey et al., 2020; Blevins et al., 2016). Furthermore, scrutinizing the implication of hip-hop in tweets can create opportunities for more robust detection of cultural signifiers in the conversations of Black Twitter subscribers (Deas et al., 2023). We hope future researchers, take care in attending to how their racialized and gendered identity influences data interpretation, and how race and gender, as fundamental causes of disease, influence group level differences in conversations online (Aguilar et al., 2023). Black women and men are more vulnerable to overdose-related deaths than their racial cohorts, and we hope this study showcases the imperative of other researchers to pay closer attention to their needs and that of Hispanic/Latinx, and American Indian populations than presently exists in drug-related research.

## Data Availability

All data produced in the present study are available upon reasonable request to the authors, provided the original public posts have not been removed by the subscribers.

## Acknowledgment

The Black women and men whose tweets provide source and inspiration for the manuscript. Coding for emotion and sentiment analysis was provided, in part, by Mohammed Al-Garadi, Research Assistant Professor at Vanderbilt University.

## Author contributions

J.W. conceptualized the study; analyzed and interpreted the data; and drafted the manuscript. A.S., S.L., and Y.G. assessed the accuracy of data analysis and reviewed code. A.S. supervised the project. Critical revision of the manuscript was completed by J.W., S.L., Y.G., and A.S.

## Statements and declarations

The authors declare no conflicting interests.

## Funding statement

Research reported in this manuscript was supported by the National Institute on Drug Abuse (NIDA) of the National Institutes of Health (NIH) under award numbers R01DA046619 and R01DA057599 (Sarker, PI) and 5T32DA050552 (Cooper, PI, Waller, PI, Sarker, PI). The content is solely the responsibility of the authors and does not necessarily represent the official views of the NIH.

## Data availability

The code for NLP will be made available upon request after publication of the manuscript. Please contact J.W. for code. Please contact A.S. or S.L. for the data.

## Ethical considerations

This study does not include human participants and informed consent is not required.

## Notes

### Competing Interest Statement

The authors have declared no competing interest.

