## supplement for "“I Been Taking Adderall Mixing it With Lean, Hope I Don’t Wake Up Out My Sleep”: Harnessing Twitter to Understand Nonmedical Prescription Stimulant Use among Black Women and Men Subscribers"

### Supplementary Tables & Figures

**S1.** Unigram Wordcloud for Women (Top 200 words)

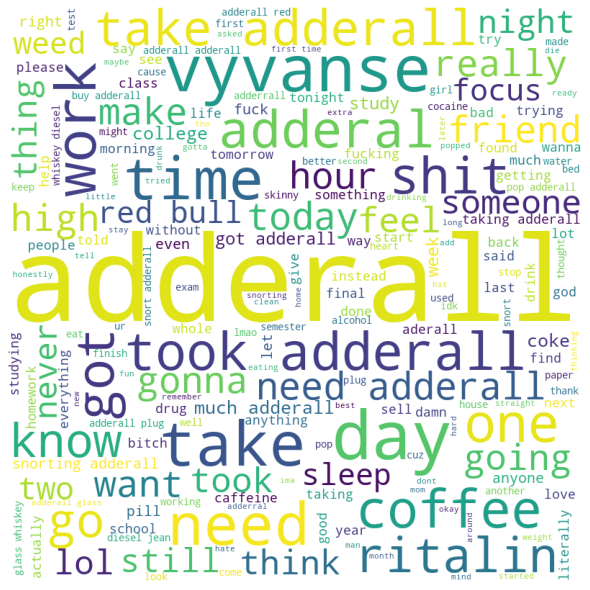

**S2.** Unigram Wordcloud for Men (Top 200 words)

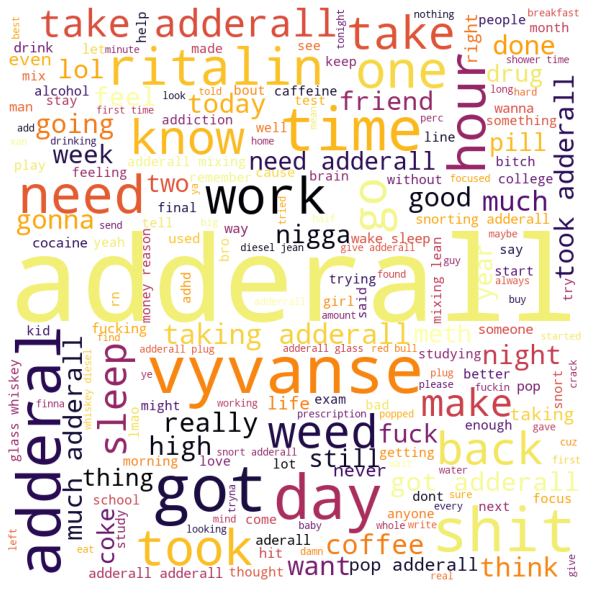

**S3.** TF-IDF Top 10 Ranked Unigrams for Women vs Men

|  | **Women** | **Men** |
| --- | --- | --- |
| **1** | adderall | adderall |
| **2** | get | like |
| **3** | take | get |
| **4** | like | got |
| **5** | need | time |
| **6** | day | take |
| **7** | took | vyvanse |
| **8** | time | day |
| **9** | vyvanse | need |
| **10** | got | took |

**S4.** TF-IDF Top 10 Ranked Bigrams for Women vs Men

|  | **Women** | **Men** |
| --- | --- | --- |
| **1** | took adderall | taking adderall |
| **2** | take adderall | take adderall |
| **3** | need adderall | took adderall |
| **4** | red bull | much adderall |
| **5** | much adderall | got adderall |
| **6** | snorting adderall | need adderall |
| **7** | get adderall | pop adderall |
| **8** | got adderall | adderall got |
| **9** | taking adderall | snorting adderall |
| **10** | adderall get | get adderall |

**S5.** TF-IDF Top 10 Ranked Trigrams for Women vs Men

|  | **Women** | **Men** |
| --- | --- | --- |
| **1** | whiskey diesel jean | taking adderall mixing |
| **2** | glass whiskey diesel | adderall mixing lean |
| **3** | adderall red bull | adderall glass whiskey |
| **4** | adderall glass whiskey | whiskey diesel jean |
| **5** | alright alright alright | glass whiskey diesel |
| **6** | time adderall glass | shower time adderall |
| **7** | shower time adderall | time adderall glass |
| **8** | adderall mixing lean | money reason gotta |
| **9** | taking adderall mixing | reason gotta take |
| **10** | day adderall red | gotta take ritalin |
